# Efficacy and side effects of the Herpes Zoster vaccine in immunocompromised individuals – a systematic review

**DOI:** 10.1101/2025.08.12.25333482

**Authors:** Daniela Marcos Raposo, David Vilhena Catarino Brito, Carla Soraia Mateus Correia, Sónia Marina Teixeira Santos, Ana Isabel Franganito Sardo

## Abstract

**Introduction:** Herpes zoster, is a reactivation of the varicella-zoster virus due to weakened immunity. It manifests as a painful rash on the skin and can become more severe in immunocompromised individuals. The risk assessment for herpes zoster among these individuals is not fully understood, but vaccination could aid in preventing this condition.

**Aim:** This study aimed to systematically review the evidence on the effectiveness and side effects of the Herpes zoster vaccine in immunocompromised patients.

**Methods:** This systematic review followed the evidence-based PRISMA protocol. The PubMed database, including Cochrane, was searched using specific MeSH terms: “herpes zoster vaccine”[MeSH Terms] AND “immunocompromised host”[MeSH Terms]. Inclusion criteria included studies published in the last 10 years involving humans of any age and gender, written in English, Spanish, French or Portuguese. Additionally, reference lists of significant studies were manually reviewed to identify relevant research that might not have been captured through electronic searches.

**Results:** The research found 10 articles. Three were considered irrelevant and removed. Out of the remaining seven, four did not meet the pre-established inclusion criteria and were excluded. Ultimately, three articles met the inclusion criteria. Additionally, five more articles from other sources were included in the study, making a total of eight eligible studies for the systematic review.

**Discussion:** The vaccination approach for preventing HZ in immunocompromised patients is controversial. A systematic literature review from 2020 found that HZ was common among all IC populations studied and exceeded the expected incidence among immunocompetent adults aged ≥60 years. The recombinant vaccine has shown high efficacy against HZ in older adults and IC populations, with all 8 studies analyzed demonstrating safety and effectiveness overall.

**Conclusion:** This overview of the efficacy of HZ prevention and vaccine immunogenicity, combined with data from previous publications, supports the use of recombinant and live attenuated HZ vaccines in the IC population. The incidence of HZ vaccine complications and their severity in IC populations generally mirrors that among those vaccinated with a placebo.

## Introduction

Herpes zoster is a reactivation of the varicella-zoster virus after it remains latent in the dorsal root and sensory nerve ganglia for decades following primary infection, which typically occurs in childhood. The precise mechanism of reactivation is not fully understood, but it is associated with a decline in cell-mediated immunity that accompanies advancing age and immunocompromised conditions such as human immunodeficiency virus infection, neoplasms, hematopoietic or solid organ transplantation, and use of immunosuppressive therapies like corticosteroids. (1–3). The disease caused by the HZ virus is usually self-limiting, with a painful rash that affects a dermatome. However, it can lead to disseminated infection in immunocompromised individuals (3).

The risk estimates for the HF population with regards to HZ have not been well characterized (2). Vaccination may be an effective strategy against HZ (1). While live, attenuated vaccines generally offer more robust immunity than inactivated virus-containing vaccines, there is a rare possibility of transmitting the vaccine virus (4). Several studies have indicated that vaccination against HZ in IC individuals can trigger immune responses and provide protection against infection (1). This study aims to systematically review the available evidence on the efficacy and adverse effects of the HZ vaccine in IC patients.

## Methods

### Characterization of the study

This systematic review was carried out based on the Preferred Reporting Items for Systematic Reviews and Meta-Analyses (PRISMA) protocol and was evidence-based. The protocol was registered in PROSPERO (ID: CRD42024498969).

### Search strategy

Scientific articles relevant to the efficacy and adverse effects of the HZ vaccine in immunocompromised patients were found through a PubMed search, including Cochrane. The search used the following MeSH terms combination: “herpes zoster vaccine”[MeSH Terms] AND “immunocompromised host”[MeSH Terms]. The inclusion criteria were: studies published in the last 10 years, involving humans of any age and gender, and written in English, Spanish, French or Portuguese. In addition to electronic searches, we manually reviewed reference lists of key studies and related articles on this topic to ensure comprehensive coverage.

### Eligibility Criteria

Titles and abstracts were assessed for relevance and study design based on predetermined inclusion criteria. Then, full articles were reviewed to identify those meeting the eligibility criteria.

Clinical trials, randomized clinical trials, systematic reviews, and meta-analyses focusing on the effectiveness and adverse effects of the HZ vaccine in IC patients were deemed eligible. Literature reviews were not included in this study.

### Ethical principles

The principles of fidelity and respect for the textual integrity of the articles were upheld by referencing the original authors when using their ideas or content. Excerpts were not used without proper context or in a different interpretative sense, in order to preserve their intended meaning.

## Results

### Study characteristics

The search in the PubMed resulted in ten articles. Three of these were not relevant to the topic and were therefore discarded. From the remaining seven articles, four did not meet the pre-established inclusion criteria and were excluded. Thus, a total of three articles from the PubMed database were included.

Five additional articles were identified and included through other sources. In total, eight studies met eligibility criteria for inclusion in this systematic review as depicted in Figure 1.

**Figure 1.**
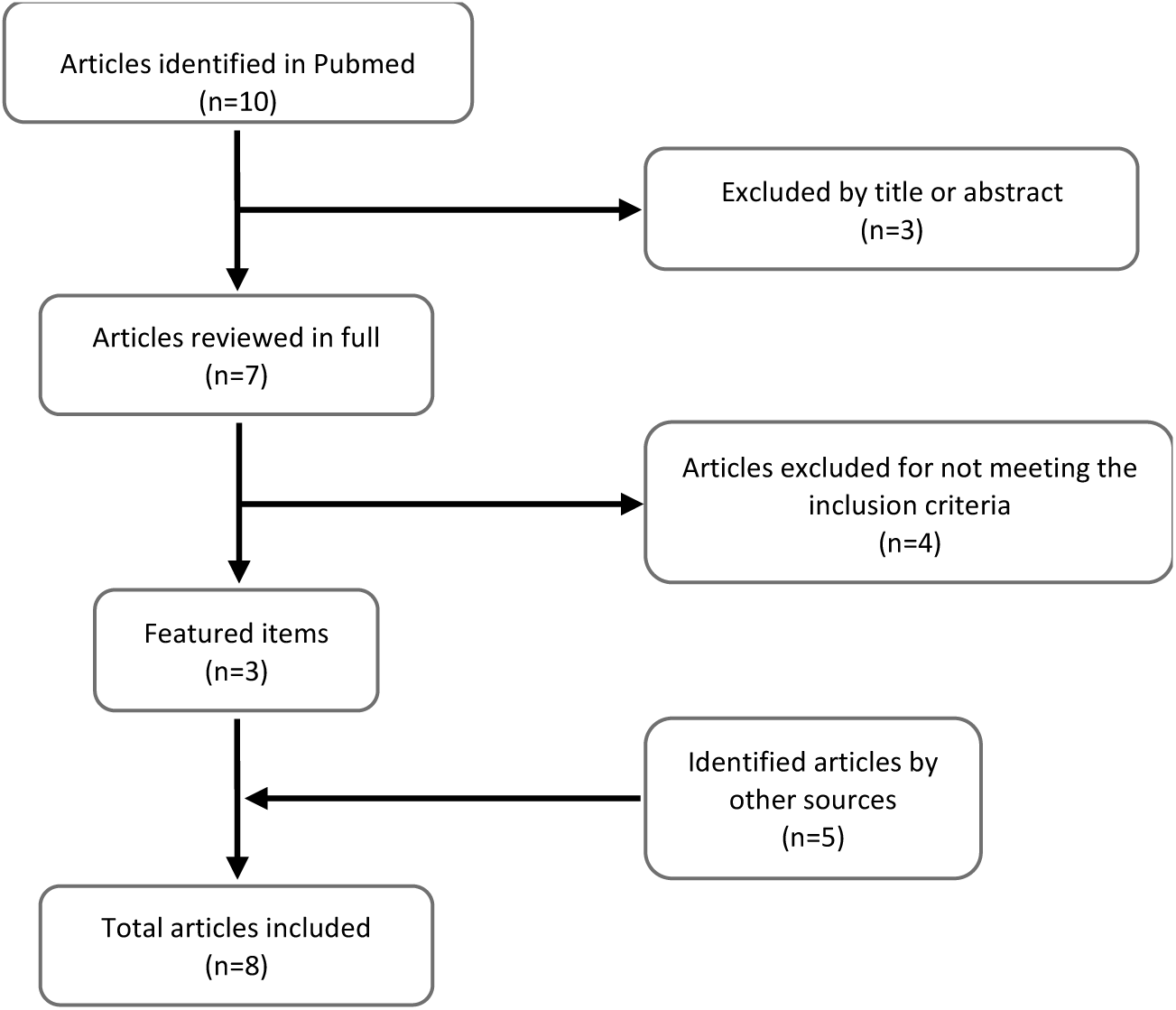
Flowchart of the research strategy and article selection: Efficacy and adverse effects of the herpes zoster vaccine in immunocompromised patients.

The main characteristics of the eligible studies are summarized in Table 1.

**Table 1.**
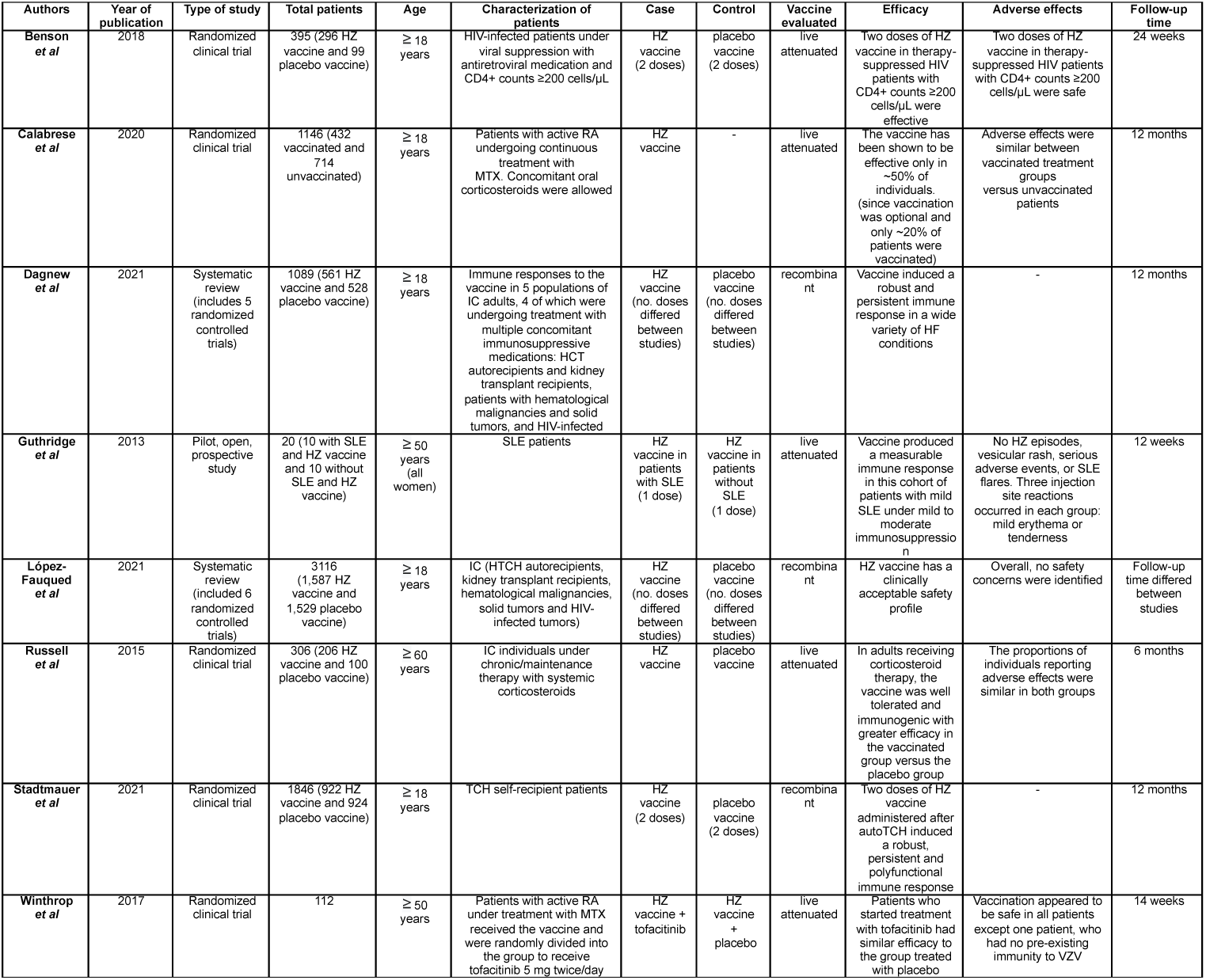
Main results of the studies analyzed.

**Table 2.** Degrees of recommendation and levels of evidence for Clinical Standards – Adapted from DGS.

| Article | Degree of recommendation | Level of evidence |
| --- | --- | --- |
| **Benson *et al*** | I | B |
| **Calabrese *et al*** | I | B |
| **Dagnew *et al*** | I | A |
| **Guthridge *et al*** | I | B |
| **López-Fauqued *et al*** | I | A |
| **Russell *et al*** | I | B |
| **Stadtmauer *et al*** | I | B |
| **Winthrop *et al*** | I | B |

### Efficacy and adverse effects of the vaccine in IC patients Recombinant vaccine

The study (5) consisted of a systematic review comparing immune responses to the recombinant HZ vaccine in 5 subpopulations of IC adults. Four of which were undergoing treatment of several concomitant immunosuppressive medications: TCH autorecipients and kidney transplants, patients with hematological malignancies, solid tumors and Human Immunodeficiency Virus (HIV) positive patients. In total, 1089 participants were involved, 91 HIV patients (54 who received the vaccine and 37 who received the placebo), 158 HCT recipient patients (82 who received the vaccine and 76 who received the placebo), 415 patients with hematological malignancies (217 who received the vaccine and 198 who received the placebo), 185 patients with solid tumors (87 who received the vaccine and 98 who received the placebo) and 214 patients undergoing kidney transplantation (121 who received the vaccine and 119 who received the placebo). In all 5 studies, participants aged ≥18 years were included. 2 doses of vaccine were administered to the different populations, except in the group of HIV patients, in which 3 doses were administered. Despite being administered in most cases when immunosuppression was close to its maximum, including concomitantly with cycles of chemotherapy, the vaccine induced a robust and persistent immune response in a wide variety of IC patient conditions, the majority of whom were under multiple immunosuppressive therapies.

Another study (6) analyzed six randomized clinical trials, which included IC participants with different clinical conditions (autologous hematopoietic stem cell transplant and kidney transplant recipients, patients with hematological malignancies, patients with solid tumors and HIV-infected adults) and placebo-controlled. The study included a total of 3116 individuals, with 1587 participants who received at least one dose of HZ recombinant vaccine were evaluated and compared with 1529 participants who received at least one dose of placebo. Of the total number of vaccine recipients, 443 (27.9%) were between 18 and 49 years old at the start of the study and 1144 (72.1%) were aged ≥ 50 at the start of the study. Of all those who received the placebo, 419 (27.4%) were between 18 and 49 years old and 1110 (72.6%) were ≥ 50 years old.

The findings indicated that the genetically engineered HZ vaccine demonstrated a safety profile with a favorable balance of benefits and risks for vaccination in immunocompromised patients. Adverse effects (AE) were monitored for 7 days following each dose as per study requirements, with additional monitoring for 30 days after vaccination for any unrequested adverse events. Serious AE, fatalities and potential immune-mediated diseases were evaluated after the first dose and up to 12 months after the last dose or end of the study. Solicited AE were more common after HZ vaccine than placebos, occurred at younger ages, and were mostly mild to moderate events. Unsolicited AE, serious AE, fatal serious AE, or potential immune-mediated illnesses were comparable between vaccinees and placebos, regardless of age group.

The study (7) consisted of a randomized clinical trial that evaluated the immunological response to the HZ recombinant vaccine in auto TCH patients. We included 1846 adult auto HTCH recipients, who were randomized to receive a first dose of vaccine (922) or placebo (924) 50-70 days after transplantation, followed by a second dose 1-2 months later. Participants were aged ≥18 and were followed for a total of 12 months. The effectiveness of the vaccine against HZ varied between 42.5% and 82.5% within the different disease groups (multiple myeloma, B and T cell non-Hodgkin lymphoma, Hodgkin lymphoma, acute myeloid leukemia and other solid neoplasms) and was statistically significant in patients with B-cell non-Hodgkin lymphoma and multiple myeloma.

To conclude, two doses of the vaccine administered after auto TCH induced a robust, persistent and polyfunctional immune response. The efficacy of the vaccine for patients with non-Hodgkin’s lymphoma was also high, despite the lower humoral response.

### Live attenuated vaccine

The research group (8) a randomized clinical trial in HIV-infected patients undergoing viral suppression with antiretrovirals. Participants, stratified by CD4+ count (200–349 or ≥350 cells/μL), were randomized to receive the HZ vaccine or placebo on day 0 and week 6. Of the 395 participants, 296 received the HZ vaccine and 99 received the placebo vaccine, aged ≥18 years and a follow-up lasting 24 weeks.

Efficacy was superior for those vaccinated versus placebos, with antibody titers being similar after 1 or 2 doses of vaccine.

Safety endpoints occurred in 15 with HZ vaccine and 2 with placebo vaccine. Injection site reactions occurred in 42% of those vaccinated vs 12.4% of placebo recipients. HZ occurred in 2 participants (1 HZ and 1 placebo), neither was related to the vaccine strain. Two doses of HZ vaccine in HIV-infected adults suppressed on therapy and with CD4+ counts ≥200 cells/μL were safe and immunogenic.

The study (9), a randomized clinical trial that evaluated patients with rheumatoid arthritis (RA) under treatment with continuous methotrexate (MTX) for ≥4 months and 15–25 mg/week for ≥6 weeks, with concomitant oral corticosteroids (≤10 mg/day of prednisolone or equivalent) were permitted. They randomly received oral monotherapy of tofacitinib 5mg BID (tofacitinib monotherapy), oral tofacitinib 5mg BID with MTX (tofacitinib plus MTX), or adalimumab (ADA) 40 mg subcutaneously every two weeks with MTX (ADA plus MTX). The study included a total of 1146 patients, 432 of whom were vaccinated, aged ≥18 years. Patients were followed up for 12 months.

Overall, the live HZ vaccine was well tolerated and adverse effects were generally similar between treatment groups and vaccinated versus unvaccinated patients. However, this study was not powered for comparisons between vaccinated and unvaccinated patients because <20% of all patients were vaccinated. Furthermore, the vaccine has only been shown to be effective in approximately 50% of individuals.

The group (10) carried out a pilot, open and prospective study that evaluated the immunological response to the live vaccine (Zostavax®) in patients with systemic lupus erythematosus (SLE). The study included 20 patients (all female), 10 with SLE and 10 controls (without disease), aged ≥50 years (SLE patients were slightly older than controls (60.5 vs. 55.3 years, respectively) who voluntarily participated in the study and were vaccinated. All participants were seropositive for VZV. Patients on immunosuppressants were included that were stable for at least 60 days before screening and considered prednisolone ≤ 10 mg/day; hydroxychloroquine ≤ 6.5 mg/kg/day; methotrexate ≤20 mg/week or azathioprine ≤150 mg/day. Patients were excluded if they had a Systemic Lupus Erythematosus Disease Activity Index (SLEDAI) >4, use of mycophenolatomofetil, cyclophosphamide, biological therapy or a daily dose of prednisolone >10 mg. The median baseline SLEDAI was 0 (range 0–2) for patients with SLE.

The results showed that vaccination produced a measurable immune response in this cohort of patients with mild SLE using mild to moderate immunosuppressants. Regarding AEs, there were no episodes of HZ, vesicular rash, serious adverse events or SLE flares. Three injection site reactions occurred in each group with mild erythema or tenderness. Follow-up occurred at 2, 6 and 12 weeks.

In the study (11), which consisted of a randomized clinical trial that evaluated the safety, tolerability and immunogenicity of the live HZ vaccine in individuals undergoing chronic/maintenance therapy with systemic corticosteroids (daily dose equivalent to 5–20mg of prednisolone) for a period ≥ 2 weeks before vaccination and ≥6 weeks after vaccination. A total of 306 participants were included (206 HZ vaccine and 100 placebo vaccine), aged ≥ 60 years, who maintained follow-up for 6 months.

In adults receiving corticosteroid therapy, the vaccine was well tolerated and immunogenic with greater efficacy in the vaccinated group versus the placebo group after vaccination.

A higher percentage of subjects reported injection site AEs in the HZ vaccine group (21.5%) compared to the placebo group (12.1%). One case of ophthalmic HZ was reported in the HZ vaccine group and considered vaccine-related by the study authors; however, Polymerase Chain Reaction testing confirmed the presence of wild-type VZV (strain not present in the vaccine).

The study (12) consisted in evaluating the efficacy and safety of the HZ vaccine in patients with active RA on MTX before starting treatment with tofacitinib 5mg twice/day or placebo 2 to 3 weeks after vaccination. A total of 112 patients were included in the study, 55 in the tofacitinib group and 57 in the placebo group, all aged ≥50 years. Follow-up took place for 14 weeks.

Patients who started treatment with tofacitinib had similar efficacy to the group treated with placebo. Adverse events occurred in 3 patients in the tofacitinib group and 0 patients in the placebo group. One patient, who had no preexisting immunity to VZV, developed cutaneous shedding of the vaccine 2 days after starting tofacitinib. Vaccination appeared to be safe in all patients except this last case.

## Discussion

Vaccination for the prevention of HZ in immunocompromised patients is still controversial, and there are no specific guidelines on its effectiveness and adverse effects of its use in this group of patients. With this systematic review we sought to clarify these questions based on the most recent scientific evidence available.

In a 2020 systematic review of the literature, which included 34 studies and estimated the risk of HZ in IC patients, it was found that HZ was common among all IC populations studied, exceeding the expected incidence of HZ among immunocompetent adults aged ≥ 60 years (2).

Vaccination has the potential to provide long-term protection against HZ, but live attenuated vaccines have been considered contraindicated in immunocompromised individuals due to the risk of developing the disease 1 s(13). On the contrary, the recombinant vaccine has demonstrated high efficacy against herpes zoster in older adults and in IC populations (14).

### Age of eligibility for vaccination

Age is a key factor when considering vaccination and according to the studies analyzed, the majority of studies involved subjects age of 18 years or more, namely the studies (5–9). In the case of studies (10, 12), participants aged 50 years or over were included and in another study, they were aged 60 years or over. In general, no study mentioned any specific reason for using these cut-offs, however, since the studies included multiple diseases and conditions that confer immunosuppression, the selection of participants was probably also carried out taking into account the age of the highest prevalence.

### Diseases and conditions that confer immunosuppression

Countless diseases require therapy that reduces immunity or cause immunosuppression themselves. In this sense, it is difficult to test all possible scenarios, and based on the review carried out, there are pathologies or treatments that, due to their greater prevalence, have more data. Rheumatological diseases, such as RA and SLE, are among the most studied groups of diseases due to their high prevalence and the need to use therapy that compromises immunity. This work included 2 studies that evaluated the HZ vaccine in patients with RA,(9) and (12) and one study that evaluated SLE (10). On the other hand, hematological and solid neoplasms, requiring immunosuppression, are a vast area of investigation; 3 studies that evaluated these conditions were included. HIV is a disease that causes immunosuppression, adding to the fact that its treatment enhances this condition, the study (8) includes patients with this pathology. Studies (5) and (11) were also included, which instead of selecting participants according to their pathologies, used medication with immunosuppressive drugs as a study inclusion criterion. This review is also in line with guidelines, which recommend that patients using methotrexate, low to moderate doses of glucocorticoids or short-term corticosteroids, intra-articular, bursa or tendon corticosteroid injections, azathioprine or 6-mercaptopurine can receive this vaccination safely (8).

Although the number of studies is limited, the most prevalent disorders were included. With this data one can, with caution, extrapolate some of the results obtained to other less studied disorders that also cause similar immunosuppression.

### Efficacy of the HZ vaccine

The effectiveness of the HZ vaccine in immunocompromised patients is a topic without official guidelines. However, due to the increased prevalence of this disease in this patient group, more research has been carried out in recent years to understand whether it can be used safely and effectively. This was one of the primary questions we aimed to address in our systematic review. Studies (8), (5), (11), (7) and (12) showed that the vaccine was more effective than those who were treated with the placebo. In the case of the study (10) that evaluated the effectiveness of the vaccine in patients with and without SLE, it was observed that the effectiveness was similar in both groups. In the study (6), it is mentioned that the vaccine has a clinically acceptable safety profile, without making particular mention of its effectiveness in the different studies it analyzed. In the study (9), since vaccination was optional and there was no placebo control group, it is difficult to extrapolate information about efficacy. In general, in the 8 studies analyzed, 2 that evaluated the recombinant HZ vaccine and another 6 that evaluated the live attenuated HZ vaccine, all of them demonstrated to be safe and effective. Quantifying the effectiveness for comparison between studies was not possible to be carried out, since different studies evaluated the effectiveness of the vaccine through the analysis of different parameters and each of them with their own particularities, particularly with regard to the inclusion criteria and exclusion, which included certain stages of severity of the disease or certain dosage of therapies used, thus making a direct comparison impossible.

### Adverse effects of the HZ vaccine

The adverse effects of the HZ vaccine are one of the concerns evaluated in immunocompromised patients. In particular, the live attenuated vaccine poses a risk of virus reactivation, especially in patients with weakened immune systems. In the studies analyzed, 6 of the 8 studies evaluated the potential risks and side effects of the HZ vaccine. Overall, they all proved to be safe, with similar proportions of adverse effects between the vaccinated groups and the placebo groups. Serious adverse effects were rare and none were associated with the vaccine.

## Conclusion

This overview of HZ prevention efficacy, vaccine immunogenicity, along with data from previous publications provides valuable medical insights and strengthens the case for using the recombinant, live attenuated HZ vaccine in the immunocompromised population at a higher risk of developing the disease. The incidence of HZ complications and their severity in IC populations generally mirrors that in those vaccinated with placebo. However, further evidence is required to shape economic and vaccination policies against HZ in this population.

## Data Availability

All data produced in the present study are available upon reasonable request to the authors

